# Associations between residential segregation, ambient air pollution, and hippocampal features in recent trauma survivors

**DOI:** 10.1101/2025.02.18.25322464

**Authors:** Sophia S. Liang, Alyssa R. Roeckner, Timothy D. Ely, Lauren A M. Lebois, Sanne J H. van Rooij, Steven E. Bruce, Tanja Jovanovic, Stacey L. House, Francesca L. Beaudoin, Xinming An, Thomas C. Neylan, Gari D. Clifford, Sarah D. Linnstaedt, Laura T. Germine, Scott L. Rauch, John P. Haran, Alan B. Storrow, Christopher Lewandowski, Paul I. Musey, Phyllis L. Hendry, Sophia Sheikh, Jose L. Pascual, Mark J. Seamon, Erica Harris, Claire Pearson, David A. Peak, Roland C. Merchant, Robert M. Domeier, Niels K. Rathlev, Brian J. O’Neil, Paulina Sergot, Leon D. Sanchez, John F. Sheridan, Steven E. Harte, Ronald C. Kessler, Karestan C. Koenen, Samuel A. McLean, Kerry J. Ressler, Jennifer S. Stevens, E. Kate Webb, Nathaniel G. Harnett

## Abstract

**Background:** Residential segregation is associated with differential exposure to air pollution. Hippocampus structure and function are highly susceptible to pollutants and associated with posttraumatic stress disorder (PTSD) development. Therefore, we investigated associations between residential segregation, air pollutants, hippocampal neurobiology, and PTSD in recent trauma survivors.

**Methods:** Participants (*N* = 278; 34% non-Hispanic white, 46% Non-Hispanic Black, 16% Hispanic) completed multimodal neuroimaging two weeks after trauma. Yearly averages of air pollutants (PM_2.5_ and NO_2_) and racial/economic segregation (Index of Concentration at the Extremes) were derived from each participant’s address. Linear models assessed if air pollutants mediated associations between segregation and hippocampal volume, threat reactivity, or parahippocampal cingulum fractional anisotropy (FA) after covarying for age, sex, income, and 2-week PTSD symptoms. Further models evaluated if pollutants or segregation prospectively predicted PTSD symptoms six months post-trauma.

**Results:** Non-Hispanic Black participants lived in neighborhoods with significantly greater segregation and air pollution compared to Hispanic and non-Hispanic white participants (*ps*<.001). There was a significant indirect effect of NO_2_ between segregation and FA values (β = 0.08, 95% CI[0.01, 0.15]), and an indirect effect of PM_2.5_ between segregation and threat reactivity (β = -0.08, 95% CI[-0.14, -0.01]). There was no direct effect of segregation on hippocampal features. Pollutants and segregation were not associated with PTSD symptoms.

**Conclusion:** Residential segregation is associated with greater air pollution exposure, which is in turn associated with variability in hippocampal features among recent trauma survivors. Further research is needed to assess relationships between other environmental factors and trauma and stress-related disorders.

## 1. INTRODUCTION

Among the 90% of Americans who will experience a traumatic event (TE) in their lifetimes, approximately 20% will subsequently develop posttraumatic stress disorder (PTSD).^1,2^ Variability in posttraumatic neurobiological and mental health outcomes may be related to individual characteristics, but it may also be influenced by environmental factors, such as levels of ambient pollution or the socioeconomic composition of the neighborhoods in which people live.^3–5^ Exposure to specific neighborhood environmental factors may modify neural circuits related to the development or expression of PTSD.^6–8^

One neighborhood characteristic implicated in the mental health literature is residential segregation, or the degree to which two or more sociodemographic groups live separately from each other.^9^ In the United States, gentrification, migration, and discriminatory real estate practices have created neighborhoods segregated along economic and racial/ethnic lines.^10^ Prior work suggests that residential segregation may be associated with differing outcomes following a TE. For example, Ahern and Galea found that six months after the September 11, 2001 terrorist attacks, neighborhood-level socioeconomic inequality was associated with depression among low-income (but not high-income) New York residents.^11^ A study of recent trauma survivors found that greater neighborhood racial and economic segregation predicted greater PTSD symptom severity.^12^ Further, several studies show that trauma-naïve individuals living in areas with greater racial and/or economic segregation report higher levels of stress, depression, and anxiety compared to individuals living in less segregated areas.^13–19^ For example, a large-scale, nationally representative survey revealed that higher Black-white metropolitan segregation was associated with higher odds of psychological distress for Black Americans beyond those associated with neighborhood poverty alone.^14^ Therefore, neighborhood segregation may be an important factor related to PTSD risk and development.

The mechanisms by which residential segregation affects posttraumatic outcomes remain unknown and are likely multifaceted, but one potential pathway may be through differential exposure to air pollution. Observational data indicate that PTSD, depression, and anxiety are more prevalent in areas with higher concentrations of air pollutants.^20–25^ Moreover, research suggests lower-income and/or ethnoracially minoritized residents disproportionately reside in neighborhoods with higher levels of hazardous air pollution caused in part by closer proximity to high-traffic routes and polluting industries.^26–29^ Exposure to two of the most common air pollutants, fine particulate matter smaller than 2.5 micrometers (PM_2.5_) and nitrogen dioxide (NO_2_), is consistently higher in areas with greater residential segregation.^27,28,30,31^ Both pollutants are products of biomass and fossil fuel combustion, particularly from automobiles, power plants, and industry,^32^ and notably, they are both associated with alterations in brain regions implicated in stress-related disorders.^23^

PM_2.5_ and NO_2_ may enter the lungs, access the brain through the bloodstream, and elevate cytokines and reactive oxygen species.^33^ The resultant neuroinflammatory reactions may damage the structure and function of regions involved in stress responding and emotion regulation, particularly the hippocampus.^33^ Preclinical models demonstrate that air pollutants can impair neurogenesis and alter morphology (e.g., decreasing dendritic spine lengths) in the hippocampus, resulting in depressive-like symptoms.^34–36^ A recent meta-analysis revealed higher levels of ambient exposure to PM_2.5_ was significantly associated with decreased hippocampal volume.^37^ Further, Dimitrov-Discher (2022) and colleagues found higher levels of PM_2.5,_ but not NO_2_, were associated with lower hippocampal reactivity in response to a social stressor.^38^ The emerging evidence on the effects of air pollutants (reviewed in Zundel et al.^23^) suggests the hippocampal alterations are similar to those implicated in PTSD.^39,40^ Altered hippocampal features associated with heightened risk for PTSD include decreased volume^40–44^, compromised white matter projections^45–46^, and lower reactivity to threat and trauma-related stimuli.^47–50^ Interestingly, several studies observed lower hippocampal volumes in individuals residing in more segregated neighborhoods, which may be due to greater exposure to air pollutants;^51–53^ however, no studies to date have investigated exposure to air pollutants as a potential mechanism by which an individual’s place of residence impacts their neurobiological risk for PTSD.

The current study investigated if trauma survivors living in more segregated neighborhoods may be exposed to higher concentrations of air pollutants and whether differential exposure to air pollutants was related to individual variability in hippocampal structure and function. We combined data from the AURORA study, a national, longitudinal study of trauma outcomes, with publicly available measures of air pollution and residential segregation. We expected that ethnoracially minoritized trauma survivors would disproportionately reside in more segregated and polluted neighborhoods compared to non-Hispanic white participants and that greater economic and racial segregation would be associated with higher levels of PM_2.5_ and NO_2_. We hypothesized that both PM_2.5_ and NO_2_ would be associated with smaller hippocampal volume, decreased white matter tract microstructure underlying the hippocampus, and lower hippocampal reactivity to threat. We further investigated whether air pollution or segregation were prospectively associated with PTSD symptoms in the six months post-trauma beyond the effects of the individual-level factors. We expected higher levels of air pollution and higher levels of segregation would be uniquely associated with more severe symptoms across time.

## 2. METHODS

### Participants

Participants were recruited as part of the AURORA study, a large-scale, longitudinal, multimodal assessment of adverse posttraumatic outcomes.^54^ The AURORA study recruited participants aged 18 to 75 from emergency departments (EDs) across the U.S. within 72 hours of experiencing a TE between September 2017 and June 2021. Qualifying TEs included motor vehicle collisions, physical assaults, sexual assaults, falls from heights greater than 10 feet, mass casualty incidents, or other events with approval of the research assistant (see ^54^ for the AURORA study’s full inclusion and exclusion criteria). Written informed consent was obtained from all participants as approved by each site’s institutional review board, and participants were financially compensated for their time.

A total of 342 participants completed the neuroimaging measures relevant to this study. Participants missing any individual characteristics were excluded (ethnoracial group *n* = 1, income *n* = 40, Week 2 PTSD symptoms *n* = 13). In addition, participants who could not be successfully geocoded and were missing neighborhood data about air pollution, neighborhood disadvantage (*n* = 9), or racial/economic segregation (*n* = 1) were excluded. The final analytic sample consisted of 278 participants. See **Figure S1** for a flowchart of inclusion/exclusion.

### Measures

#### Demographics

Sociodemographic data was collected in the ED (at the time of study enrollment) and at two weeks post-trauma. Participants provided their current home address, which was used for geocoding. In the ED, participants reported their sex assigned at birth (*male* or *female*), age, education, and race and ethnicity (queried separately). The race and ethnicity responses were combined to create an ethnoracial group variable with the following categories: non-Hispanic white (“NH white”), non-Hispanic Black (“NH Black”), Hispanic, and non-Hispanic Other (“NH Other”). At the two-week study visit, participants gave an estimate of their annual household income before taxes on a semi-continuous scale (≤*$19,000*, *$19,001–$35,000*, *$35,001–$50,000*, *$50,001–$75,000*, *$75,001–$100,000*, or *>$100,000*).

#### Psychometric Assessments

At two weeks, eight weeks, three months, and six months post-trauma, PTSD symptoms were assessed using the PTSD Checklist for DSM-5 (PCL-5). The two-week survey asked participants to assess the symptoms they experienced within the last two weeks, while the eight-week survey onwards asked participants to reference the past 30 days. Potential scores range from 0 to 80, where higher scores corresponded to more severe symptoms.^55^ In addition, depression and anxiety symptoms were assessed using subsets of questions from the Patient-Reported Outcomes Measurement Information System (PROMIS).^56^ The depression variables from Short Form 8b were converted into a T-score, where a score of 50 is the mean for the U.S. general population and a score of 60 or above (at least one standard deviation above the mean) indicates a suspect depression diagnosis. Anxiety symptoms were assessed using the PROMIS Anxiety Bank. Scores ranged from 0 to 16, where a higher score is indicative of more severe anxiety.

### Neighborhood-level Factors

#### Residential Segregation

Residential segregation was measured using the Index of Concentration at the Extremes (ICE), a metric developed by Massey, Booth and Crouter ^57^ and expanded upon by Krieger, Waterman, Gryparis and Coull ^26^. The ICE measures the extent to which the population in a certain area is concentrated into extremes of relative privilege and deprivation. The ICE ranges from -1 to +1, where -1 indicates that 100% of the population in an area is concentrated in the most deprived group, and +1 indicates that 100% of the population is concentrated in the most privileged group. The two groups may be defined along any socioeconomic axis, and a growing body of literature shows that the ICE is a reliable predictor of physical and mental health disparities among individuals segregated by income levels and ethnoracial categories.^12,58–61^. Other scholars have also noted that a conceptual strength of the ICE, as opposed to more traditional metrics such as neighborhood poverty rate, is that it reflects the zero-sum relationship between privilege and deprivation — emphasizing the fact that certain groups have gained and maintained their advantages *at the expense of* others.^62^

Population, income, and race/ethnicity data for each zip code tabulation area in the U.S. were obtained from the U.S. Census Bureau’s American Community Survey for 2016, the year before the AURORA study began recruitment. These data were used to compute various ICE metrics using code from Krieger et al (https://www.hsph.harvard.edu/thegeocodingproject/covid-19-resources/). For the present analyses, we used the combined ethnoracial and economic segregation measure (ICEraceinc; herein referred to as residential segregation), which defines the privileged group as NH white Americans at or above the 80^th^ percentile of national family income and the deprived group as people of color at or below the 20^th^ percentile of national family income.

#### Air Pollutants

Air pollution measures included the estimated annual concentrations of PM_2.5_ and NO_2_ for each zip code in the U.S.^63^ The estimations reflect the ensemble predictions of several machine-learning models using data from air monitoring, satellites, meteorological conditions, chemical transport models, and land-use variables. These estimates have high predictive performance and have been widely used to study the effects of air pollutants on population health.^64^ The present study used the PM_2.5_ and NO_2_ measures from 2016.

#### Neighborhood Socioeconomic Disadvantage

Several previous studies using the AURORA data have quantified neighborhood advantage/disadvantage using the Area Deprivation Index (ADI), a weighted-composite measure of income, education, employment, and housing quality.^7,65^ Therefore, as a secondary analysis, we replaced the ICEraceinc with ADI rankings in statistical tests to determine if our results were specific to segregation. We used the 2019 ADI, based on data from the 2015-2019 censuses. The ADI of each census block group is ranked against all other block groups nationwide and converted into a percentile (1–100), such that a national ADI ranking of 100 would indicate that a block group was the most socioeconomically disadvantaged of all block groups in the U.S.

### MRI Data Acquisition and Analysis

The AURORA study collected MRI data at two weeks post-trauma at five sites nationwide with largely harmonized acquisition protocols; site-specific sequences are provided in Supplemental Table S1.^66,67^ Structural, diffusion weighted, and functional images were visually inspected and underwent quality checks in MRI-QC.^68^ Additional information about the MRI quality control metrics considered for each modality is reported in the Supplemental Material.

#### Structural MRI

T1-weighted MRI data were analyzed using Freesurfer as part of fMRIPrep.^66^ Each individual’s hippocampal volume was standardized as a proportion of their total intracranial volume.

#### Diffusion Tensor Imaging (DTI)

White matter microstructure was assessed using DTI and processed according to the recommendations of the ENIGMA consortium, as described in prior work.^66^ Standard ENIGMA skeletal maps of fractional anisotropy (FA) were first projected on each participant’s FA maps before extracting FA values from the Johns Hopkins University white matter atlas. The tract of interest in this study was the parahippocampal part of the cingulum.

#### Task-fMRI

As reported elsewhere^66^, participants completed a task viewing fearful and neutral facial expressions while undergoing fMRI. In each 8 s block, participants viewed 8 different faces (presented for 500 ms with a 500 ms interstimulus interval) depicting either fear or neutral expressions. In all, the task consisted of 15 fearful and 15 neutral blocks presented in a pseudorandom order, counterbalanced across participants. As described elsewhere, data were preprocessed in fMRIPrep, and 1^st^-level models were conducted in SPM12. Separate boxcar functions were used to represent each block, convolved with a canonical hemodynamic response function, and separate regressors for white matter, cerebrospinal fluid, and global signal were included to account for any remaining noise. The current analysis examined hippocampus reactivity to *fearful > neutral* faces.

### Analytic Strategy

We completed Pearson’s correlation analyses to examine associations between the individual, neighborhood, air pollution, and hippocampal continuous variables. One-way ANOVAs examined ethnoracial group differences in income and education, ICEraceinc, ADI, PM_2.5_, NO_2_, hippocampal volume, CGH FA values, and hippocampal threat reactivity respectively. A mediation analysis was conducted in R (*process* package, Model 4 ^69^) with bias-corrected bootstrapping (20,000 resamples) to examine the relationship between residential segregation on each of the neurobiological measures of the hippocampus (volume, CGH FA, threat reactivity) via an association with air pollution (PM_2.5_ or NO_2_ concentrations). In line with prior work,^37^ to isolate the relative importance of neighborhood-level factors on hippocampal features from the effects of individual-level sociodemographic and trauma-related factors, individual sex, age, income, and Week 2 PTSD symptoms were included as covariates. Due to collinearity between neighborhood characteristics and neuroimaging site, site was not included as a covariate in these analyses. Ninety-five percent confidence intervals were estimated for all these effects; intervals that excluded the value 0 were considered statistically significant. A secondary analysis examined whether associations held when using ADI as the independent variable instead of ICEraceinc (see Supplemental Material). Finally, to examine whether segregation or air pollution predicted PTSD development over time, we conducted a linear mixed effects model analyzing the effects of ICEraceinc, NO_2_, and PM_2.5_ on PTSD symptoms across all four timepoints, after adjusting for sex, age, and income. As an exploratory aim, analyses of highly comorbid depression and anxiety symptoms were conducted, and results are provided in (see Supplemental Material).

## 3. RESULTS

### Sample Characteristics and Associations Between Study Measures

Of the 278 participants, the majority had an annual household income of under $35,000, and a plurality identified as NH Black (46%). Over two-thirds of participants experienced a motor vehicle collision. See **Table 1** for a breakdown of participant demographics.

**Table 1.**
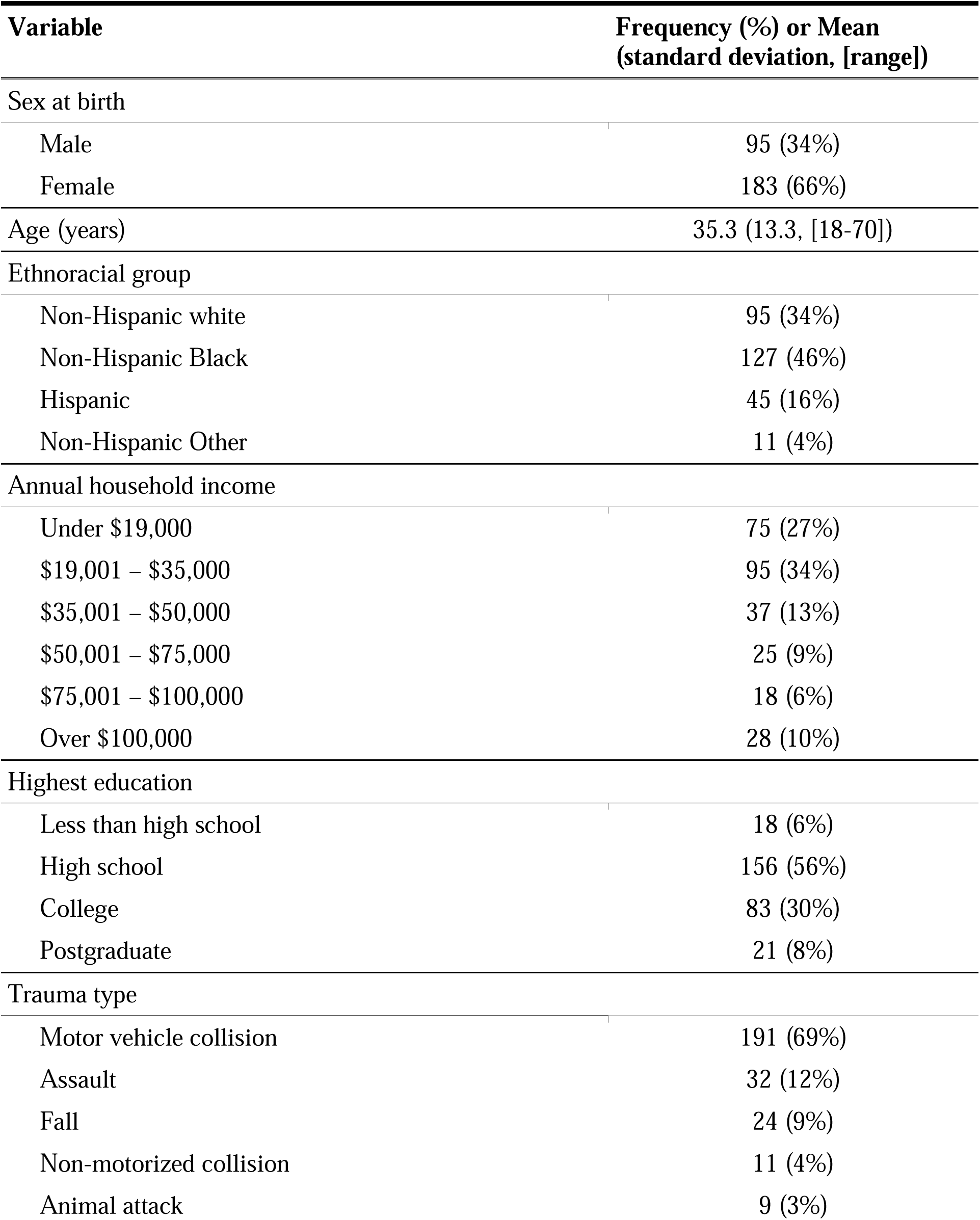
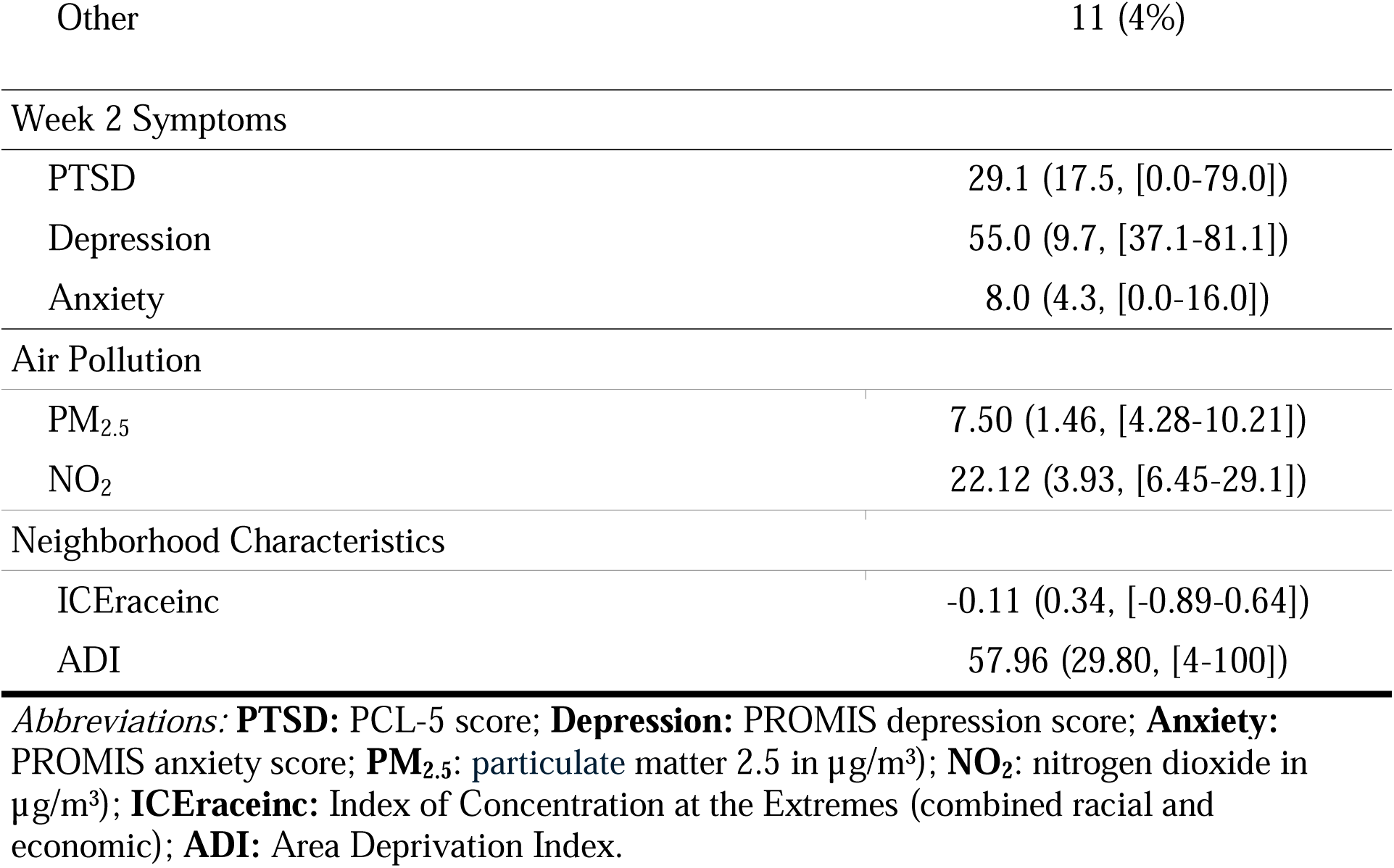
Sample characteristics.

| **Variable** | **Frequency (%) or Mean**<br>**(standard deviation, [range])** |
| --- | --- |
| Sex at birth |  |
| Male | 95 (34%) |
| Female | 183 (66%) |
| Age (years) | 35.3 (13.3, [18-70]) |
| Ethnoracial group |  |
| Non-Hispanic white | 95 (34%) |
| Non-Hispanic Black | 127 (46%) |
| Hispanic | 45 (16%) |
| Non-Hispanic Other | 11 (4%) |
| Annual household income |  |
| Under \$19,000 | 75 (27%) |
| \$19,001 – \$35,000 | 95 (34%) |
| \$35,001 – \$50,000 | 37 (13%) |
| \$50,001 – \$75,000 | 25 (9%) |
| \$75,001 – \$100,000 | 18 (6%) |
| Over \$100,000 | 28 (10%) |
| Highest education |  |
| Less than high school | 18 (6%) |
| High school | 156 (56%) |
| College | 83 (30%) |
| Postgraduate | 21 (8%) |
| Trauma type |  |
| Motor vehicle collision | 191 (69%) |
| Assault | 32 (12%) |
| Fall | 24 (9%) |
| Non-motorized collision | 11 (4%) |
| Animal attack | 9 (3%) |
| Other | 11 (4%) |
| Week 2 Symptoms |  |
| PTSD | 29.1 (17.5, [0.0-79.0]) |
| Depression | 55.0 (9.7, [37.1-81.1]) |
| Anxiety | 8.0 (4.3, [0.0-16.0]) |
| Air Pollution |  |
| PM<sub>2.5</sub> | 7.50 (1.46, [4.28-10.21]) |
| NO<sub>2</sub> | 22.12 (3.93, [6.45-29.1]) |
| Neighborhood Characteristics |  |
| ICERaceinc | -0.11 (0.34, [-0.89-0.64]) |
| ADI | 57.96 (29.80, [4-100]) |

*Abbreviations:* **PTSD:** PCL-5 score; **Depression:** PROMIS depression score; **Anxiety:** PROMIS anxiety score; **PM<sub>2.5</sub>:** particulate matter 2.5 in $\mu\text{g}/\text{m}^3$ ; **NO<sub>2</sub>:** nitrogen dioxide in $\mu\text{g}/\text{m}^3$ ; **ICERaceinc:** Index of Concentration at the Extremes (combined racial and economic); **ADI:** Area Deprivation Index.

There was a significant main effect of ethnoracial group on residential segregation (*F*(3, 274) = 39.4, *p* < .001), exposure to PM_2.5_ (*F*(3, 274) = 17.3, *p* < .001), and exposure to NO_2_ (*F*(3, 274) = 7.29, *p* < .001) (**Figure 1**). Tukey post hoc tests revealed these effects were largely driven by NH Black participants, who lived in more segregated and disadvantaged neighborhoods (a more negative ICEraceinc) compared to Hispanic (difference in means = -0.21; *t*(274) = 4.30, *p* < .001), NH white (diff = -0.41; *t*(274) = 10.79, *p* < .001), and NH Other (diff = -0.28; *t*(274) = - 3.12, *p* = .011) participants, *p*s < .001. Hispanic participants lived in more segregated and disadvantaged neighborhoods compared to NH white participants (diff = -0.20; *p* < .001; *t*(274) = -3.97, *p* = .001). NH Black participants were exposed to higher concentrations of PM_2.5_ compared to Hispanic (diff = 1.21; *t*(274) = 4.30, *p* < .001), NH white (diff = 1.14; *t*(274) = - 6.28, *p* < .001), and NH Other participants (diff = 1.14; *t*(274) = 2.69, *p* = .038), as well as higher concentrations of NO_2_ compared to NH white participants (diff = 2.25; *p* < .05; *t*(274) = -4.35, *p* < .001.

**Figure 1.**
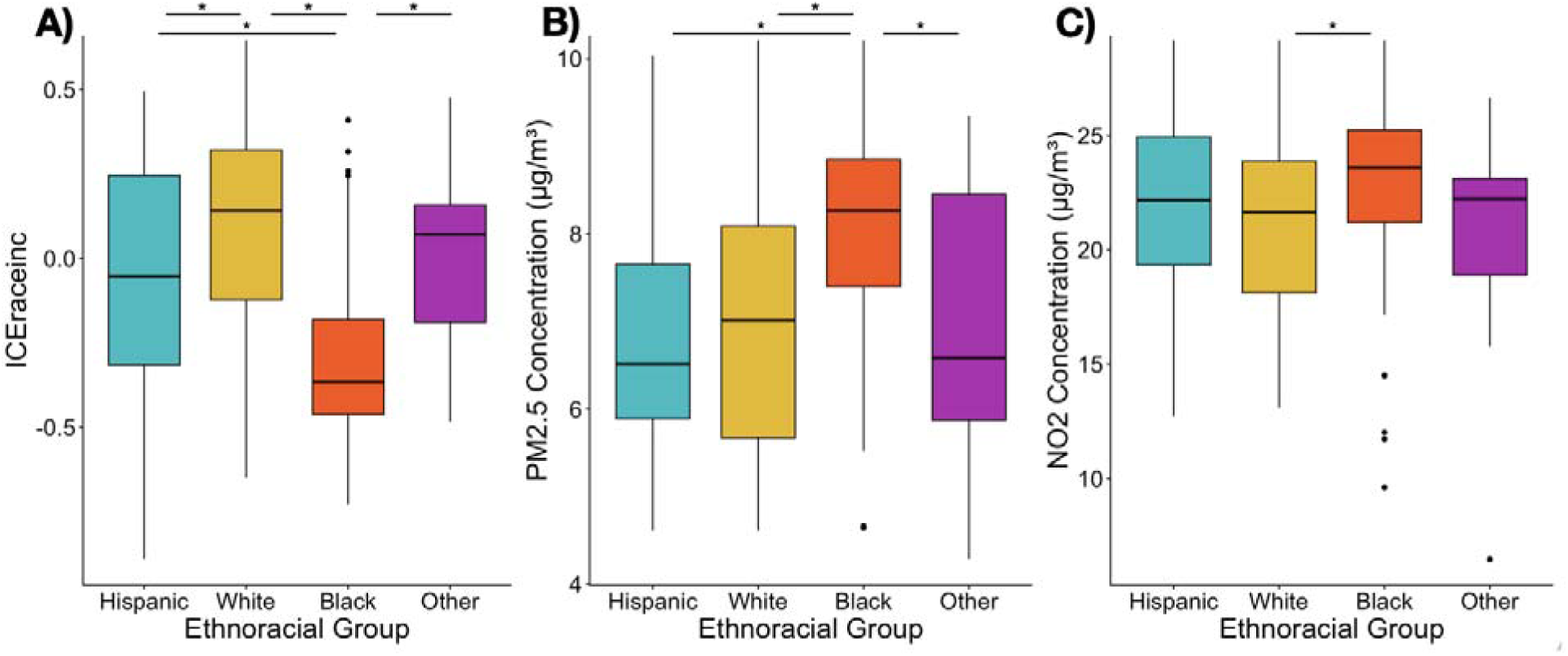
Ethnoracial differences between residential segregation and air pollutant exposure. There was a significant main effect of ethnoracial group on residential segregation, exposure to PM_2.5_, and exposure to NO_2_ (* represents *p* < .05 between each pair straddled by horizontal lines). **[A]** Non-Hispanic Black participants lived in more segregated and disadvantaged neighborhoods (a more negative ICEraceinc) compared to Hispanic, non-Hispanic white, and non-Hispanic Other participants. Hispanic participants lived in more segregated and disadvantaged neighborhoods compared to non-Hispanic white participants. **[B]** Non-Hispanic Black participants were exposed to higher concentrations of PM_2.5_ compared to Hispanic, non-Hispanic white, and non-Hispanic Other participants. **[C]** Non-Hispanic Black participants were exposed to higher concentrations of NO_2_ compared to non-Hispanic white participants.

Scatter plots showing the relationships of air pollution concentration with bilateral hippocampus volume, white matter tract FA, and threat reactivity are depicted in **Figure 2**. PM_2.5_ concentration was positively correlated with threat reactivity (*r*(276) = 0.16, *p* < .006). NO_2_ concentration was positively correlated with hippocampus volume (*r*(276) = 0.17, *p* < .005) and negatively correlated with white matter tract FA (*r*(276) = -0.18, *p* < .003).

**Figure 2.**
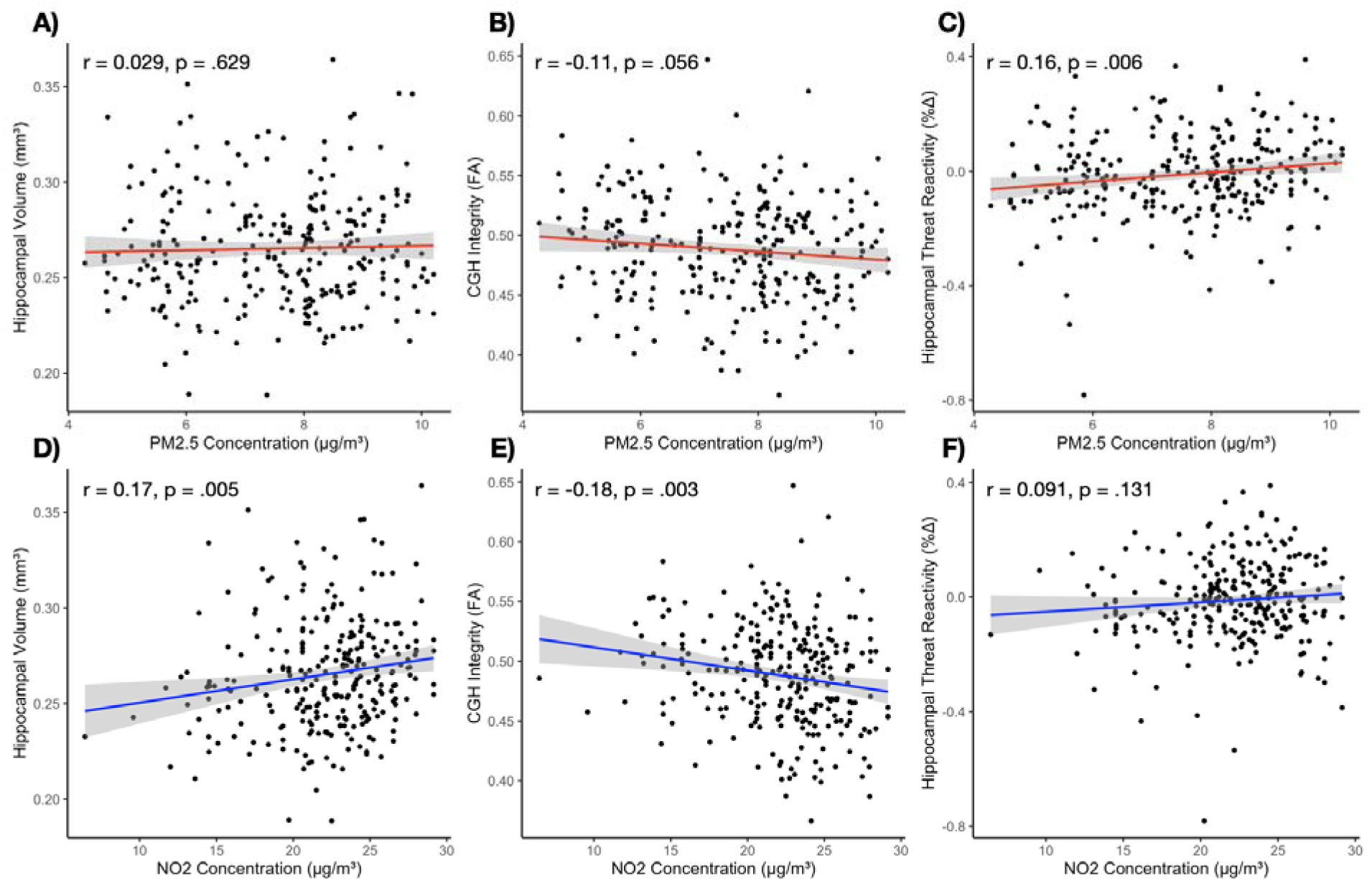
Bivariate associations between air pollutants and hippocampal features. PM_2.5_ concentration was not associated with **[A]** hippocampus volume or **[B]** parahippocampal cingulum (CGH) tract integrity as measured by fractional anisotropy (FA); however, it was **[C]** positively correlated with hippocampal threat reactivity (*p* < .05). NO_2_ concentration was **[D]** positively correlated with hippocampal volume and **[E]** negatively correlated with CGH FA values (*p*s < .05). **[F]** NO_2_ was not associated with hippocampal threat reactivity. (Dots represent individual participants, lines represent the linear lines of best fit, and shading represents the 95% confidence interval.)

### Residential Segregation, Air Pollution, and Hippocampal Features

A mediation model revealed that there was a significant indirect association between segregation, NO_2_ exposure, and FA of the CGH (a*b path: β = 0.08, CI [0.01, 0.15], SE = 0.04; **Figure 3A**) covarying for sex, age, income, and Week 2 PCL-5 scores. Greater segregation was associated with greater NO_2_ exposure, which in turn was associated with lower FA values of the CGH. There was no significant direct association between segregation and the CGH (c’ path: β = 0.01, CI [-0.01, 0.02], SE = 0.01).

**Figure 3.**
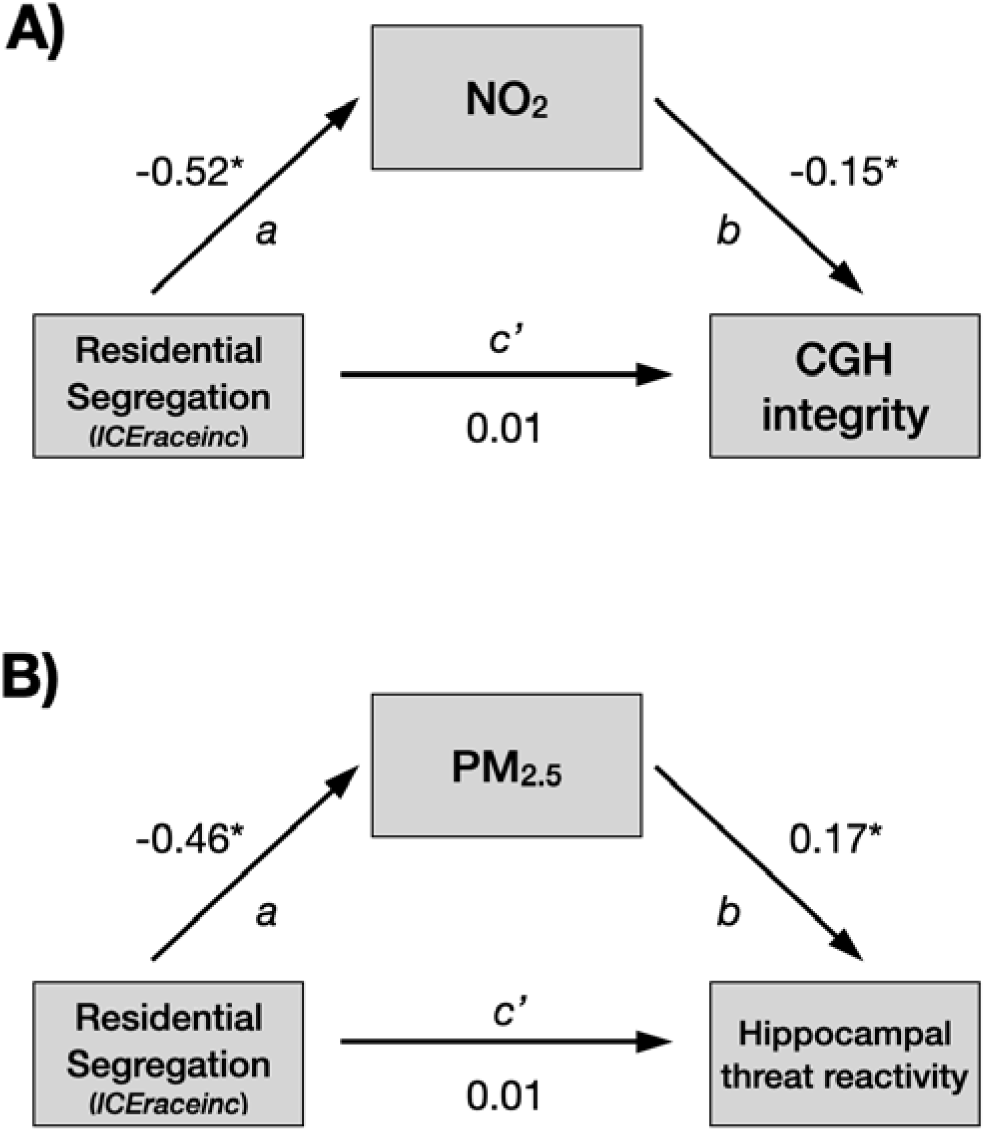
Path analysis of associations between residential segregation, air pollutants, and hippocampal features. **[A]** After covarying for sex, age, income, and Week 2 PCL-5 scores, NO_2_ was a significant mediator of the effect of residential segregation (ICEraceinc) on the integrity of the parahippocampal cingulum (CGH) (a*b path: β = 0.08, CI [0.01, 0.15], SE = 0.04). There was no direct association between residential segregation and the CGH (c’ path: β = 0.01, CI [-0.01, 0.02], SE = 0.01). **[B]** PM_2.5_ was a significant mediator of the effect of residential segregation on threat reactivity (a*b path: β = -0.08, CI [-0.14, - 0.01], SE = 0.033). There was no direct association between residential segregation and threat reactivity (c’ path: β = 0.01, CI [-0.05, 0.07], SE = 0.03). (* represents a value outside of the 95% confidence interval)

There was a significant indirect association between segregation, PM_2.5_ exposure, and threat reactivity (a*b path: β = -0.08, CI [-0.14, -0.01], SE = 0.03; **Figure 3B**); greater segregation was associated with greater PM_2.5_ exposure, which in turn was associated with increased threat reactivity. There was no significant direct association between segregation and threat reactivity (c’ path: β = 0.01, CI [-0.05, 0.07], SE = 0.03). When ICEraceinc was replaced with ADI, there were no significant associations between ADI and threat reactivity, but there was an indirect association between ADI, NO_2_, and volume, as well as ADI, NO_2_, and FA of the CGH (see Supplemental Material).

### Residential Segregation, Air Pollution, and PTSD Symptoms

There was no significant association between residential segregation and PTSD symptoms (β = 0.01, *p* > .05; **Table 2**) after covarying for sex, age, and income. Further, there was no significant association between either PM_2.5_ (β = -0.01, *p* > .05) or NO_2_ (β = -0.06, *p* > .05) on PTSD symptoms across the four timepoints (**Table 2**). Follow-up exploratory models of depression and anxiety similarly did not reveal associations between symptoms and residential segregation (**Tables S2 and S3**).

**Table 2.** Results from the linear mixed effect model predicting PTSD symptoms with air pollutants and residential segregation.

| Predictors | Estimates | Std. Beta | Standardized Std. Error | Standardized CI | p-value |
| --- | --- | --- | --- | --- | --- |
| *Intercept* | 34.39 | 0.01 | 0.05 | -0.09 – 0.11 | <.001 |
| PM<sub>2.5</sub> Concentration (µg/m<sup>3</sup>) | -0.08 | -0.01 | 0.06 | -0.13 – 0.12 | .920 |
| NO<sub>2</sub> Concentration (µg/m<sup>3</sup>) | -0.31 | -0.06 | 0.06 | -0.19 – 0.06 | .315 |
| ICERaceinc | 0.76 | 0.01 | 0.06 | -0.11 – 0.14 | .829 |
| Time | -2.66 | -0.17 | 0.02 | -0.20 – -0.13 | <.001* |
| Sex [*female*] | 6.04 | 0.16 | 0.05 | 0.06 – 0.26 | .002* |
| Age (yrs) | 0.11 | 0.08 | 0.05 | -0.02 – 0.18 | .128 |
| Annual Family Income (\$) | -1.97 | -0.18 | 0.06 | -0.29 – -0.06 | .002* |

**Abbreviations:** **ICERaceinc:** Index of Concentration at the Extremes (combined racial and economic segregation); **PM<sub>2.5</sub>:** particulate matter 2.5; **NO<sub>2</sub>:** nitrogen dioxide. *Note:* \* indicates $p < .05$

## 4. DISCUSSION

The present study explored a potential pathway by which social and spatial inequities may be associated with posttraumatic neurobiology. We found significant associations among residential segregation, air pollution, and hippocampal features in recent trauma survivors across the United States even after accounting for important individual-level factors. Higher residential segregation was associated with higher levels of NO_2_ and PM_2.5_, to which NH Black participants were disproportionately exposed. Further, higher levels of air pollution were associated with decreased white matter tract integrity and increased threat reactivity in the hippocampus. Our findings suggest that individuals’ environments may impact hippocampal neurobiology in trauma survivors and replicate well-documented economic and ethnoracial inequities in air pollution exposure.^70,71^

Our results align with previous preclinical and human work suggesting air pollutants are associated with altered neurobiological features.^23,72^ The majority of prior work has focused on the hippocampus and PM ^23^ with relatively few studies characterizing the unique effects of multiple pollutants, partly due to the complexities of disentangling pollutant mixtures and the high collinearity between pollutant types^23^. In the present study, we found that higher levels of NO_2_ were associated with lower FA values of the CGH, a tract that facilitates communication between the hippocampus and prefrontal regions, whereas higher levels of PM_2.5_ were associated with greater hippocampal reactivity to threat. Several studies have documented NO_2_ and PM_2.5_ are associated with reduced microstructural integrity, with the majority showing childhood exposure is associated with lower global fractional anisotropy later in life.^73–75^ Relatively few human studies have examined associations between air pollutants and threat or stress reactivity. Interestingly, Dimitrov-Discher (2022) and colleagues found higher levels of PM_2.5,_ but not NO_2_, were associated with *lower* hippocampal responses to a social stressor. However, Miller and colleagues (2019) demonstrated that adolescents exposed to higher levels of PM_2.5_ showed greater psychophysiological reactivity during a stress test as evidenced by lower heart rate variability and higher skin conductance levels.^76^ Together, these studies emphasize that higher levels of air pollutants are related to neurobiological changes that may be associated with altered threat and stress responding.

There is currently no established pattern that NO_2_ or PM_2.5_ preferentially affect microstructure or function. Indeed, both PM_2.5_ and NO_2_ can penetrate the lungs and initiate an immune response. The resulting systemic inflammation may lead to structural and functional changes; with rodent research indicating hippocampal changes (e.g., reductions in dendritic spine length) can occur within hours or days of exposure.^23^ Differential effects of various pollutants may be observed due to differences in pollutant size, exposure duration, lag times, or the biological mechanisms targeted (e.g., necrosis, neuroinflammation, alterations to neurotransmitters or their receptors).^23^ For example, larger particulate matter (PM_10_) may enter the lungs, but PM_2.5_ is small enough to be absorbed in the blood stream.^32^ A recent meta-analysis of neuroimaging work suggested higher levels of ambient exposure to PM_2.5_ were more strongly associated with decreased hippocampal volume compared to PM_10_ or NO_2_.^37^ However, exposure durations are not always accurately reported, which may influence how robustly specific pollutants are associated with neurobiological or psychiatric outcomes. In general, long-term pollutant exposure appears more closely associated with macro- and micro-structural changes over the life course whereas short-term exposure is highly correlated with neural activity.^73^ Further research is needed to understand the unique and interactive impact of different air pollutant concentrations and exposure times on the hippocampus and the central nervous system more broadly.

In the context of recent trauma exposure, the implications of alterations of these features are mixed. For example, several studies among trauma-exposed individuals indicate that lesser CGH integrity is associated with more severe PTSD^45,77^ while others suggest lower FA values are associated with less severe PTSD symptoms.^78,79^ Inconsistencies also exist in the relationship between hippocampal reactivity to threat and PTSD. Prior AURORA study analyses reveal an inverse relationship between hippocampal reactivity to fearful faces and PTSD symptoms.^47^ However, youth with PTSD show greater hippocampal reactivity to threat compared to youth without PTSD and those with remitting PTSD.^80^ More work is therefore needed to determine how or whether the neurobiological patterns associated with air pollution exposure are related to PTSD risk. The patterns of results may also be more related to psychiatric outcomes unmeasured in this study. For example, lesser CGH integrity is more consistently associated with other psychiatric outcomes, including mild cognitive impairment and Alzheimer’s disease.^81^ Finally, preclinical work suggests air pollutants are associated with poor performance on hippocampus-dependent tasks, including impaired contextual fear memory.^82^ The present study only examined hippocampus activity during a passive threat task, and future work should also consider investigating the relationship between air pollutants and hippocampal activity during a fear learning and memory paradigm.

Notably, while we found that residential segregation was significantly associated with higher levels of air pollutants and subsequently differences in hippocampal structure and function, residential segregation and NO_2_ or PM_2.5_ were not significantly associated with symptoms of PTSD. It is possible that the effects of pollution exposure on PTSD symptoms may take longer than six months to manifest, which would be missed in our acutely traumatized sample. Further, there may be underreporting of symptoms from NH Black participants, who live in the most segregated and polluted neighborhoods in this sample; stigma surrounding mental illness is more common among racially minoritized groups and may contribute to underreporting in this study.^83^ Alternatively, NH Black participants, who disproportionately experience chronic socioeconomic and/or environmental stressors, may develop adaptive responses to future stressors, buffering the harmful effects of the TE.^84^ Finally, another possible explanation is that the present measurement of air pollutants did not fully capture each individual’s personal exposure level. The yearly average was derived from models using data from regional air quality monitors, and the number of monitoring stations varies substantially across American cities. Air pollutant exposure measured with personal and mobile monitors may be more closely associated with an individual’s mental health, and future work should consider both personal and neighborhood exposure.^85^

### Limitations

The present findings should be interpreted in light of several limitations. Home address was collected at study enrollment, and residential stability or length of time at residence was not assessed. It is unknown if participants lived in the same neighborhood, neighborhoods similar in composition, or significantly different neighborhoods throughout their lifespan. Further, it was not possible to determine the chronicity of exposure or characterize any fluctuations in residential segregation or pollution exposure that may have important neuropsychiatric effects. However, prior work suggests long-term pollutant exposure is more closely associated with structural changes while short-term exposure is highly correlated with neural activity.^73^ Future studies may therefore expand on our work to investigate both individual exposure and environmental changes within a neighborhood over time. Additionally, we did not account for any physical health conditions participants may have had. Although the AURORA study surveyed participants’ assessments of their overall health and asked whether their health prevented them from engaging in physical activities, it did not collect information about specific diagnoses. Both residential segregation and exposure to air pollution are associated with adverse physical health outcomes such as asthma and hypertension,^34,62^ and these conditions may affect hippocampal features^34,86^ and posttraumatic symptoms in ways unaccounted for in our work. Further research is also needed to examine nuances within residential segregation itself, as there may be social and cultural factors associated with living in segregated, homogenous neighborhoods that are *beneficial* to mental health, such as closer ties with neighbors.^87^ Some studies of predominantly Asian or Hispanic neighborhoods, for example, have found that recent immigrants can find greater social capital and cultural resources within ethnic enclaves that offset socioeconomic disadvantages and bolster their mental health.^5,59,87,88^

### Conclusions

The present study highlights important relationships between socioenvironmental factors and hippocampal neurobiology. We found that NH Black trauma survivors were exposed to disproportionately higher levels of air pollution. Air pollution, in turn, was associated with variability in hippocampal structure and function, which have been heavily implicated in PTSD. Future research that integrates assessments of societal-level factors may offer a more nuanced understanding of neurobiological correlates of trauma responses, which may help guide early identification of individuals most at-risk in the aftermath of a TE. Such work may also guide policy efforts to address these environmental inequities across American cities and determine potential long-term effects of inequities on health.^89^

## Data Availability

Data used in this manuscript is available through the National Institute of Mental Health (NIMH) Data Archive (NDA). The NDA Collection for the AURORA Project can be found here: https://nda.nih.gov/edit_collection.html?id=2526. This content is solely the responsibility of the authors and may not reflect the official view of any of the funders or the Submitters submitting original data to NDA.

## Acknowledgements

The investigators wish to thank the trauma survivors participating in the AURORA Study. Their time and effort during a challenging period of their lives make our efforts to improve recovery for future trauma survivors possible. The AURORA study was supported by NIMH under U01MH110925, the US Army MRMC, One Mind, and The Mayday Fund. The DISENTANGLE study is a continuation of AURORA’s work and is supported by the United States Army Medical Research Acquisition Activity (USAMRAA) under Contract No. W81XWH22C012. This project was supported by NIMH under F32MH134443 (Webb) and K01MH129828 (Harnett), Data used in this manuscript is available through the National Institute of Mental Health (NIMH) Data Archive (NDA). The NDA Collection for the AURORA Project can be found here: https://nda.nih.gov/edit_collection.html?id=2526. This content is solely the responsibility of the authors and may not reflect the official view of any of the funders or the Submitters submitting original data to NDA.

