## Supplemental Material for "Associations between residential segregation, ambient air pollution, and hippocampal features in recent trauma survivors"

**Supplemental Methods**

Figure S1 provides a study flowchart and reasons participants were excluded in the current analyses. Table S1 describes the harmonized MRI acquisition parameters across each site.

***MRI Quality Control and Exclusions***

The MRIQC pipeline (version 0.10.4 in a Docker container) (Esteban et al. 2017) was run on structural and functional images for quality control. For T1-weighted (T1w) images, we evaluated the coefficient of joint variation (CJV) of gray and white matter. CJV is thought to reflect the presence of heavy head motion and artifacts, where lower scores reflect better quality data. T1w images with a CJV greater than 2 standard deviations from the mean (by imaging site) were excluded. For functional data, we assessed frame-wise displacement (FD). Functional images with greater than 15% of volumes exceeding 1mm FD were excluded from analyses. For diffusion tensor images, we visually inspected initial DWI data for the appearance of technical artifacts or heavy motion that would impact preprocessing, and participants with such artifacts were excluded.

A total of 412 participants had useable structural MRI (n = *7* were excluded for anatomical reasons; *n* = 6 were excluded for excessive motion (based on CJV); *n* = 3 were excluded for technical issues). Of these 412 participants, 6 did not complete the threat reactivity task, 14 were excluded due to missing behavioral data, 17 were excluded due to excessive motion, and 9 were excluded due to technical issues, resulting in 366 participants with useable T1w and threat reactivity data. Of these participants, 8 did not complete diffusion tensor imaging, 6 were excluded based on motion criteria, and 10 were excluded due to technical issues. Therefore, a total of 342 participants had usable data across all modalities.

**Supplemental Results**

In the mediation model where ICE was replaced with the Area Deprivation Index (ADI), there was a significant indirect association between ADI, NO_2_, and hippocampal volume (a*b path: β = 0.04, CI [0.00, 0.08], SE = 0.02); greater neighborhood disadvantage was associated with greater NO_2_ exposure, which was in turn associated with greater hippocampal volume (Supplemental Figure 2A). There was no significant direct association between ADI and volume (c’ path: β = 0.00, CI [0.00, 0.00], SE = 0.00).

There was also a significant indirect association between ADI, NO_2_, and FA of the CGH (a*b path: β = -0.05, CI [-0.10, -0.01], SE = 0.02); greater neighborhood disadvantage was associated with greater NO_2_ exposure, which was in turn associated with decreased FA in the CGH (Supplemental Figure 2B). There was no significant direct association between ADI and CGH integrity (c’ path: β = 0.00, CI [0.00, 0.00], SE = 0.00).

In the linear mixed models, after controlling for individual characteristics (sex, age, income), there was no significant association between air pollution and depression or anxiety symptoms. Time, sex, and annual family income predicted depression and anxiety symptoms, and age predicted anxiety symptoms, *p*s < .05 (Tables S2 and S3). However, neither PM_2.5_ nor NO_2_ were significant predictors of depression or anxiety symptoms across the four timepoints, *p*s > .05.

**Table S1.** Harmonized MRI Sequences Across Study Sites

|  | Site 1 | Site 2 | Site 3 | Site 4 | Site 5 |
| --- | --- | --- | --- | --- | --- |
| Scanner | Siemens TIM 3T Trio | Siemens TIM 3T Trio | Siemens MAGNETOM 3T Prisma | Siemens 3T Verio | Siemens MAGNETOM 3T Prisma |
| Head Coil | 12 Channel | 12 Channel | 20 Channel | 12 Channel | 20 Channel |
| Modality |  |  |  |  |  |
| T1-weighted | **TR** = 2530ms, **TEs** = 1.74/3.6/5.46/7.32ms, **TI** = 1260ms, **flip angle** = 7, **FOV** = 256mm, **slices** = 176, **Voxel size** = 1mm x 1mm x 1mm | **TR** = 2530ms, **TEs** = 1.74/3.6/5.46/7.32ms, **TI** = 1260ms, **flip angle** = 7, **FOV** = 256mm, **slices** = 176, **Voxel size** = 1mm x 1mm x 1mm | **TR** = 2300ms, **TE** = 2.96ms, **TI** = 900ms, **flip angle** = 9, **FOV** = 256mm, **slices** = 176, **Voxel size** = 1.2mm x 1.0mm x 1.0mm | **TR** = 2530ms, **TEs** = 1.79/3.65/5.51/7.37ms, **TI** = 1260ms, **flip angle** = 7, **FOV** = 256mm, **slices** = 176, **Voxel size** = 1mm x 1mm x 1mm | **TR** = 2530ms, **TEs** = 2.24/4.1/5.96/7.82ms, **TI** = 1350ms, **flip angle** = 7, **FOV** = 256mm, **slices** = 176, **Voxel size** = 1mm x 1mm x 1mm |
| Diffusion Weighted Imaging | **TR** = 7700ms, **TE** = 85ms, **FOV** = 212mm, **flip angle** = 90, **Volumes** = 71 (64 **b**=1000 s/mm^2,^ 7 b0), **PA-encoded**, **Voxel size** = 2mm x 2mm x 2mm | **TR** = 7700ms, **TE** = 85ms, **FOV** = 212mm, **flip angle** = 90, **Volumes** = 71 (64 **b**=1000 s/mm^2,^ 7 b0), **PA-encoded**, **Voxel size** = 2mm x 2mm x 2mm | **TR** = 7000ms, **TE** = 74ms, **FOV** = 212mm, **flip angle** = 90, **Volumes** = 71 (64 **b**=1000 s/mm^2,^ 7 b0), **PA-encoded**, **Voxel size** = 2mm x 2mm x 2mm | **TR** = 12000ms, **TE** = 85ms, **FOV** = 212mm, **flip angle** = 90, **Volumes** = 71 (64 **b**=1000 s/mm^2,^ 7 b0), **PA-encoded**, **Voxel size** = 2mm x 2mm x 2mm | **TR** = 7700ms, **TE** = 67ms, **FOV** = 212mm, **flip angle** = 90, **Volumes** = 71 (64 **b**=1000 s/mm^2,^ 7 b0), **PA-encoded**, **Voxel size** = 2mm x 2mm x 2mm |
| fMRI | **TR** = 2360ms, **TE** = 30ms, **flip angle** = 70, **FOV** = 210mm, **slices** = 44, **Voxel size** = 3mm x 3mm x 3mm, 0.5 mm gap | **TR** = 2360ms, **TE** = 30ms, **flip angle** = 70, **FOV** = 210mm, **slices** = 44, **Voxel size** = 3mm x 3mm x 3mm, 0.5 mm gap | **TR** = 2360ms, **TE** = 29ms, **flip angle** = 70, **FOV** = 210mm, **slices** = 44, **Voxel size** = 3mm x 3mm x 3mm, 0.5 mm gap | **TR** = 2360ms, **TE** = 30ms, **flip angle** = 70, **FOV** = 210mm, **slices** = 42, **Voxel size** = 3mm x 3mm x 3mm, 0.5 mm gap | **TR** = 2360ms, **TE** = 29ms, **flip angle** = 90, **FOV** = 210mm, **slices** = 44, **Voxel size** = 3mm x 3mm x 3mm, 0.5 mm gap |

**Table S2.** Results from the linear mixed effect model predicting depression symptoms with air pollutants and segregation (* indicates *p* <.05)

| **Predictors** | **Estimates** | **Std. Beta** | **Standardized Std. Error** | **Standardized CI** | ***p*-value** |
| --- | --- | --- | --- | --- | --- |
| *Intercept* | 62.29 | 0.00 | 0.05 | -0.10 – 0.10 | <.001 |
| PM_2.5_ Concentration (µg/m³) | -0.44 | -0.06 | 0.06 | -0.19 – 0.06 | .336 |
| NO_2_ Concentration (µg/m³) | -0.17 | -0.06 | 0.06 | -0.19 – 0.06 | .332 |
| ICEraceinc | 1.59 | 0.05 | 0.06 | -0.07 – 0.18 | .419 |
| Time | -0.94 | -0.10 | 0.02 | -0.14 – -0.06 | <.001* |
| Sex [*female*] | 2.23 | 0.10 | 0.05 | 0.00 – 0.20 | .042* |
| Age (yrs) | 0.06 | 0.08 | 0.05 | -0.02 – 0.18 | .111 |
| Annual Family Income ($) | -1.42 | -0.22 | 0.06 | -0.33 – -0.11 | <.001* |

σ^2^ = 37.02

τ_00 PID_ = 62.16

ICC = 0.63

N_PID_ = 278

Observations = 984

Marginal R^2^ / Conditional R^2^ = 0.073 / 0.644

*Abbreviations:* **ICEraceinc:** Index of Concentration at the Extremes (combined racial and economic segregation); **PM_2.5_**: particulate matter 2.5; **NO_2_**: nitrogen dioxide. *Note:* * indicates *p* <.05

**Table S3.** Results from the linear mixed effect model predicting anxiety symptoms with air pollutants and residential segregation (* indicates *p* <.05)

| **Predictors** | **Estimates** | **Std. Beta** | **Standardized Std. Error** | **Standardized CI** | ***p*-value** |
| --- | --- | --- | --- | --- | --- |
| *Intercept* | 9.63 | 0.00 | 0.05 | -0.10 – 0.09 | <0.001 |
| PM_2.5_ Concentration (µg/m³) | -0.12 | -0.04 | 0.06 | -0.16 – 0.08 | .523 |
| NO_2_ Concentration (µg/m³) | -0.09 | -0.07 | 0.06 | -0.20 – 0.05 | .226 |
| ICEraceinc | 0.59 | 0.04 | 0.06 | -0.08 – 0.17 | .472 |
| Time | -0.62 | -0.16 | 0.02 | -0.19 – -0.12 | <0.001* |
| Sex [*female*] | 1.64 | 0.17 | 0.05 | 0.08 – 0.27 | <0.001* |
| Age (yrs) | 0.04 | 0.12 | 0.05 | 0.02 – 0.22 | .015* |
| Annual Family Income ($) | -0.52 | -0.19 | 0.05 | -0.30 – -0.08 | .001* |

σ^2^ = 7.24

τ_00 PID_ = 10.60

ICC = 0.59

N_PID_ = 278

Observations = 981

Marginal R^2^ / Conditional R^2^ = 0.102 / 0.636

*Abbreviations:* **ICEraceinc:** Index of Concentration at the Extremes (combined racial and economic segregation); **PM_2.5_**: particulate matter 2.5; **NO_2_**: nitrogen dioxide. *Note:* * indicates *p* <.05

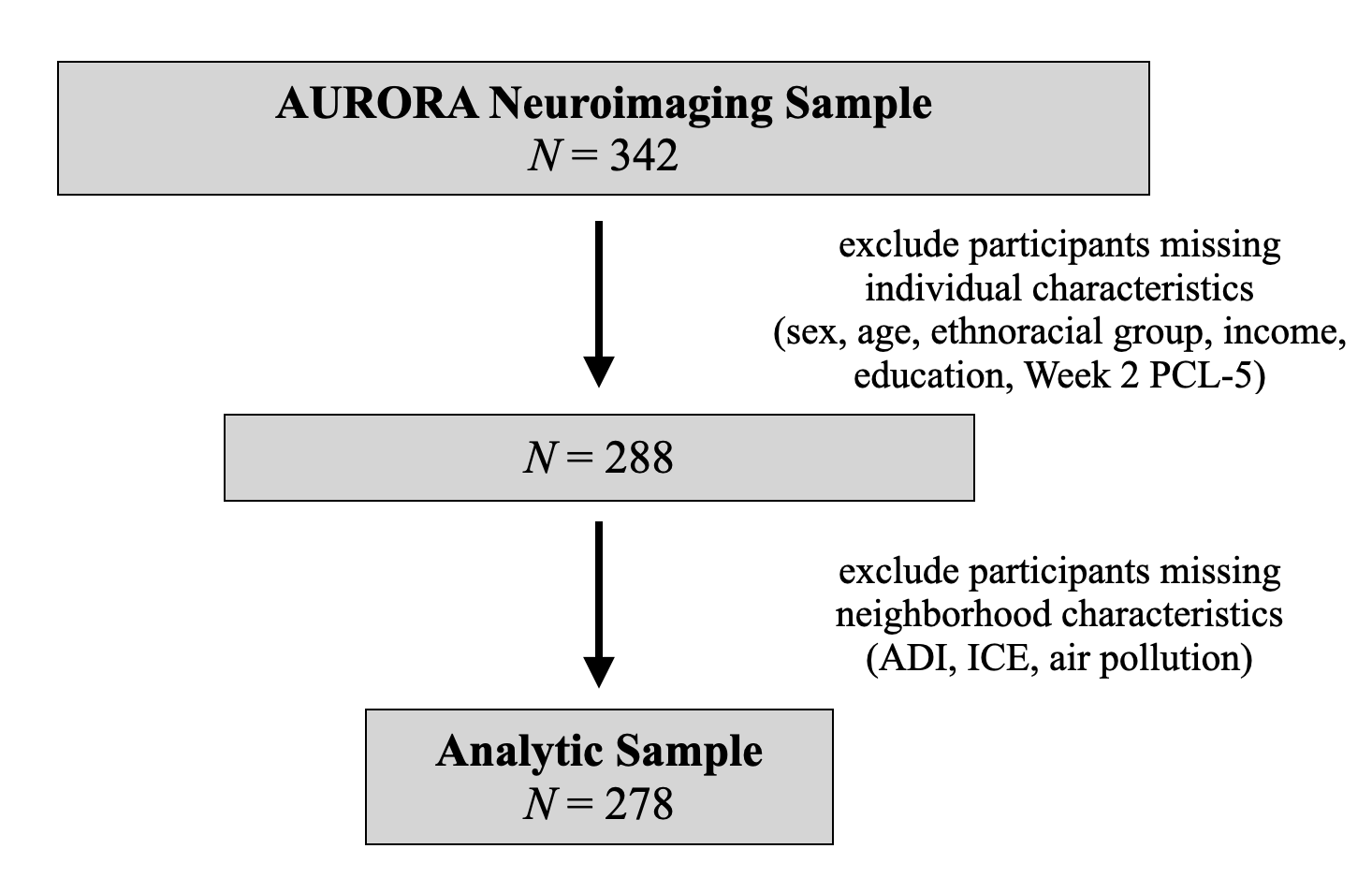

**Supplemental Figure 1.** Flowchart of inclusion/exclusion criteria. After participants missing individual or neighborhood characteristics were excluded, there were 278 participants remaining in the analytic sample.

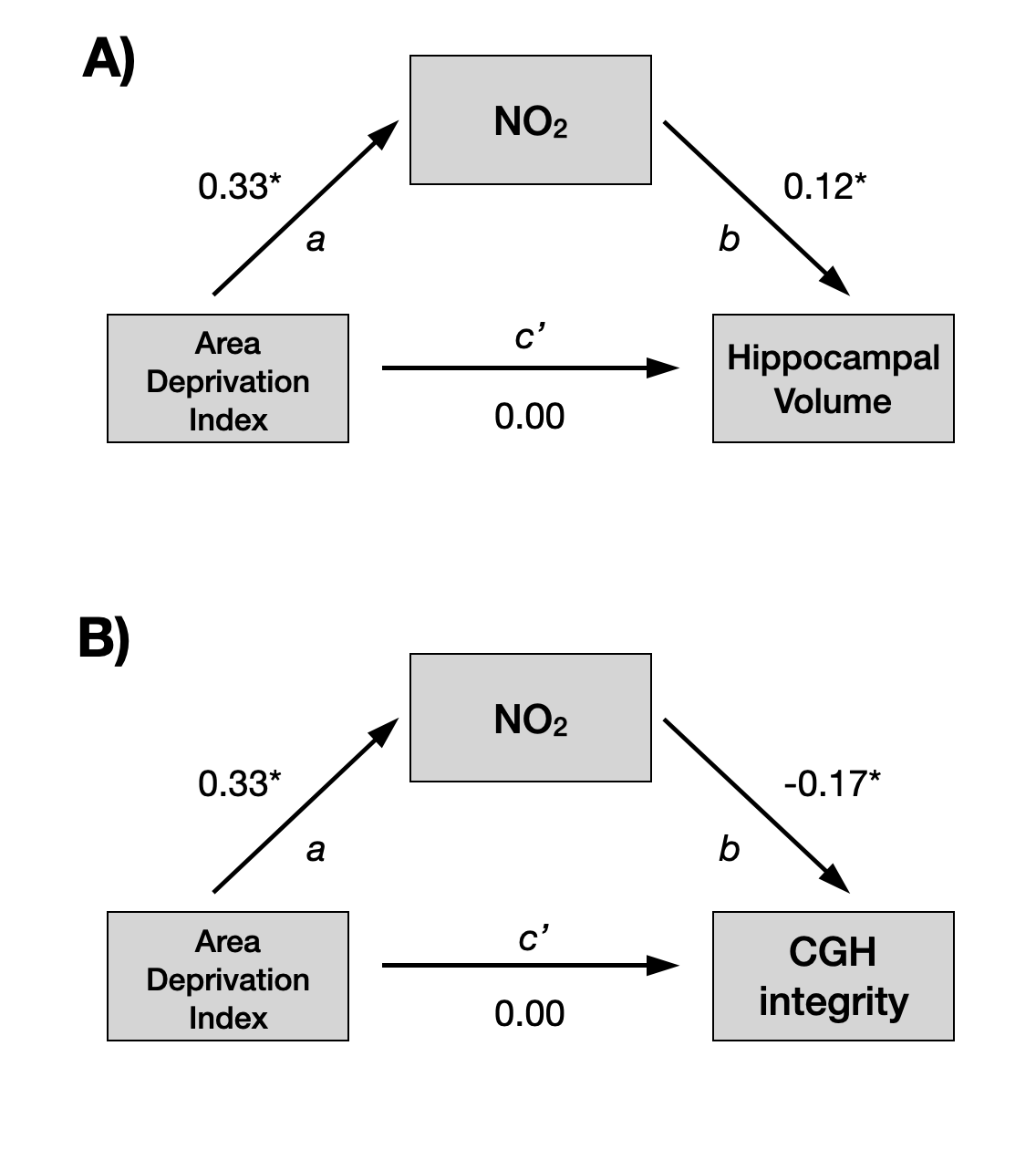

**Supplemental Figure 2. [A]** After covarying for sex, age, income, and Week 2 PCL-5 scores, NO_2_ was a significant mediator of the effect of neighborhood deprivation (ADI) on hippocampal volume (a*b path: β = 0.04, CI [0.00, 0.08], SE = 0.02). There was no significant direct association between ADI and volume (c’ path: β = 0.00, CI [0.00, 0.00], SE = 0.00). **[B]** NO_2_ was a significant mediator of the effect of neighborhood deprivation on the integrity of the parahippocampal cingulum (CGH) (a*b path: β = -0.05, CI [-0.10, -0.01], SE = 0.02). There was no significant direct association between ADI and CGH integrity (c’ path: β = 0.00, CI [0.00, 0.00], SE = 0.00). (* represents a value outside of the 95% confidence interval).
